# A consideration of publication-derived immune-related associations in Coronavirus and related lung damaging diseases

**DOI:** 10.1101/2020.04.15.20065425

**Authors:** Nophar Geifman, Anthony D. Whetton

## Abstract

The severe acute respiratory syndrome virus SARS-CoV-2, a close relative of the SARS-CoV virus, is the cause of recent COVID-19 pandemic affecting, to date, nearly 2 million individuals across the globe and demonstrating relatively high rates of infection and mortality. A third virus, the H5N1, responsible for avian influenza, has caused infection with some clinical similarities to those in COVID-19 infections.

Cytokines, small proteins that modulate immune responses, have been directly implicated in some of the severe responses seen in COVID-19 patients, e.g. cytokine storms.

Understanding the immune processes related to COVID-19, and other similar infections, could help identify diagnostic markers and therapeutic targets.

Here we examine data of cytokine, immune cell types, and disease associations captured from biomedical literature associated with coronavirus, SARS, and H5N1 influenza, with the objective of identifying potentially useful relationships and areas for future research. Cytokine and cell-type associations captured from MeSH terms linked to thousands of PubMed abstracts, has identified differing patterns of associations between the three corpuses of abstracts (coronavirus, SARS, or H5N1 influenza). Clustering of cytokine-disease co-occurrences in the context of coronavirus has identified compelling clusters of co-morbidities and symptoms, some of which already known to be linked to COVID-19. Finally, network analysis identified sub-networks of cytokines and immune cell types associated with different manifestations, co-morbidities and symptoms of coronavirus, SARS, and H5N1.

Systematic review of research in medicine is essential to facilitate evidence-based choices about health interventions. In a fast moving pandemic the approach taken here will identify trends and enable rapid comparison to the literature of related diseases.

## INTRODUCTION

The recent respiratory disease pandemic (COVID-19) [1], caused by the severe acute respiratory syndrome (SARS) coronavirus 2 (SARS-CoV-2), has both high rates of infection as well as high mortality rates [2, 3]. In many inflammatory diseases, signaling proteins termed cytokines play a critical role in disease pathology. These proteins are secreted mainly from hematopoietic cells including lymphocytes and macrophages, and along with such cells, play a central role in many diseases as well as in health [4-7]. In the case of COVID-19, and other viral diseases, they can significantly affect the course and outcome for the patient. This may manifest itself as a cytokine release syndrome or cytokine storm, observed in many patients as highly elevated levels of these proteins during acute disease. While first linked to the 1918 influenza pandemic [8] the cytokine storm was extensively documented in the avian H5N1 influenza virus infection [9]. H5N1 influenza virus causes acute lung injury as also observed in COVID-19. Acute respiratory distress syndrome (ARDS) is the principal cause of respiratory failure associated with severe influenza as it is with COVID-19 and other members of the Coronavirus family. In severe influenza infections the extent of lung injury is due to dysregulated inflammatory responses. Hence, to compare and contrast immune-related trends in H5N1 influenza and coronavirus infections could enable clinical research to more effectively consider treatment options.

SARS-CoV-2 is closely related to SARS-CoV, the virus responsible for the 2003 SARS pandemic. Since COVID-19 is an emerging pandemic, there is currently less data available on the involvement of the immune system. However, the resource available for Coronaviruses more generally, SARS more specifically, as well as H5N1 influenza, makes it possible to gain insight into the pathology associated with SARS-CoV-2, in respect of pro-inflammatory events and lung damage.

One rich, readily available source of immune- and disease-related knowledge is the corpus of published scientific research. Within research publications, a copious amount of disease-related trends are captured; these can be freely extracted from PubMed records [10]. We have previously developed a framework for extracting disease-immune relationships from PubMed abstracts by relying on the linking of such abstracts to concepts from Medical Subject Headings (MeSH) [11]. MeSH is the National Library of Medicine’s controlled vocabulary thesaurus, consisting of terms (naming and descriptors) within a hierarchical structure; they are used for indexing MEDLINE PubMed publications. MeSH descriptors associated with each MEDLINE citation are manually assigned and provide a straightforward, and useful, knowledge resource. Numerous works using concept co-occurrences in biomedical texts or in associated MeSH terms have shown the utility of MeSH in capturing biomedical knowledge [12-18]. Our own assessment of the ability of MeSH descriptors associated within the same PubMed record to represent a true (meaningful and feasible) relationship between the terms has shown that co-occurrence of MeSH descriptors linked to any given PubMed record are a good source for mining dependencies between different types of biomedical entities [11].

The work presented here examines immune-related molecular and cellular patterns in the context of Coronaviruses, SARS, and in H5N1 influenza. By identifying links between these conditions and different immune system players, novel research can be better targeted at areas of greater impact.

## METHODS

### Extracting PubMed Record-Associated MeSH Descriptors

A complete description of the approach taken by this study can be found in [11]. Briefly, a list of cytokine MeSH descriptors (such as “interferon gamma,” “transforming growth factor beta,” and “chemokine CCL3”) was manually compiled by a domain expert, who browsed MeSH’s sub-trees and selected those descriptors that were deemed relevant to this work.

Similarly, a list of immune-related cell type MeSH descriptors (such as “lymphocytes,” “Th1 cells,” and “basophils”) was also compiled (see Supplementary File 1). A comprehensive list of disease names and synonyms was extracted from the Human Disease Ontology. This approach to capturing associations between disease, cytokines and different cell types had been extensively evaluated, as described in [11]. For each PubMed abstract (see below) the associated MeSH descriptors were recorded, and the lists of cytokine names, cell types, and diseases were searched for exact matches.

### PubMed data

The PubMed database was searched on April 6^th^, 2020 for abstracts tagged with either the ‘Severe Acute Respiratory Syndrome’ MeSH term, the ‘influenza a virus, h5n1 subtype’ MeSH term, or with the ‘Coronavirus’ MeSH term (excluding those with the SARS Mesh term). These searches formed the SARS corpus with a total of 4493 abstracts, the H5N1 corpus with a total of 5975 abstract, and the Coronavirus corpus with a total of 9810 abstracts, respectively (Figure 1).

**Figure 1:**
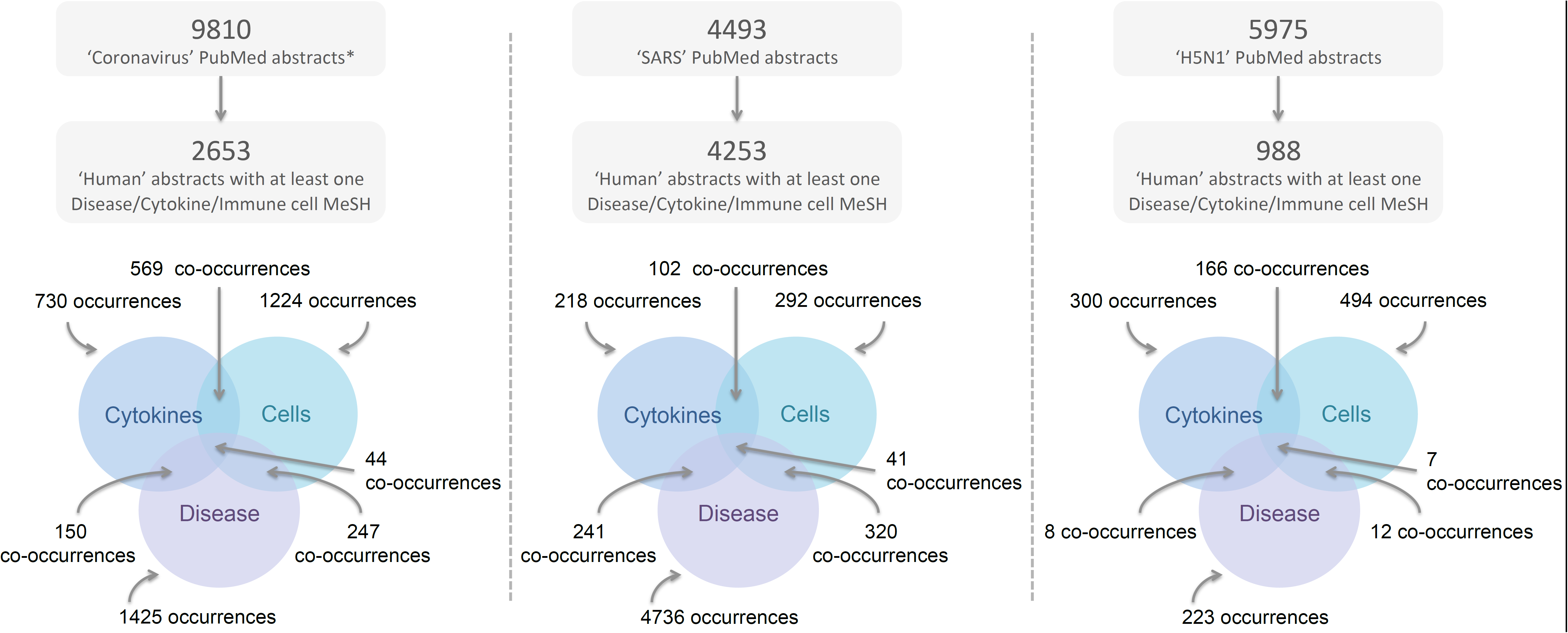
Extraction of PubMed record-associated MeSH descriptors. The lower Venn diagrams represents the occurrences of the different types of MeSH descriptors (disease, cell, or cytokine), with overlapping areas representing the co-occurrences between the different term types. * PubMed was searched for abstracts linked to the ‘Coronavirus’ MeSH term but not to the SARS MeSH term.

### Clustering and correlation analysis

In order to examine disease similarities based on cytokine or cell-type co-occurrences in the context of Coronavirus or SARS, we set out to cluster these patterns of co-occurrences in the literature. To do so, a quantitative cytokine-disease or cell-disease matrix was generated by obtaining, for each disease and cytokine/cell in the data, a count of the number of records mapped to that disease and that cytokine/cell.

Using these matrices as input, hierarchical clustering was performed using (1-correlation) as the distance measure.

### Network analysis

Network analysis was carried out with the Cytoscape software. One network was created using the information extracted from the Coronavirus (but not SARS) corpus; a second network was created using data extract from the SARS corpus; and a third was created for H5N1. Sub-networks were created by selecting nodes (primarily disease nodes) of interest and then selecting first-degree neighbors. Node and edge attributes such as, node type, node counts (of occurrences) and edge counts (of co-occurrences) were used for colouring, sizing, and layout of the networks.

## RESULTS

Using a previously published approach (Geifman et al 2015) associations between cytokines, immune cell types, and diseases were captured from PubMed records related to Coronavirus, SARS and H5N1 influenza. Differing patterns of associations were identified in a corpus of over 9000 Coronavirus (excluding SARS) related abstracts and compared to those obtained in a corpus of over 4000 SARS related abstracts, and a corpus of nearly 6000 H5N1 related abstracts.

### Different cytokines and cell types are associated with Coronavirus, SARS and H5N1 influenza

In a comparison of the number of associations of cytokines and immune cells with abstracts between the three corpuses, some interesting differences were identified (Figure 2 and Supplementary File 1). For example, Coronavirus is more highly associated with Interferon-gamma than both SARS and H5N1; while Interferon-beta is less associated with SARS-related abstracts than Coronavirus- or H5N1-related abstracts (Figure 2A). On the other hand, SARS -related abstracts have a higher percentage of those linked to CXCL10 (interferon-γ inducible protein 10) and Interleukin (IL)-8 (Figure 2A). Other differences between the three corpuses are found for different immune cells (Figure 2B).

**Figure 2:**
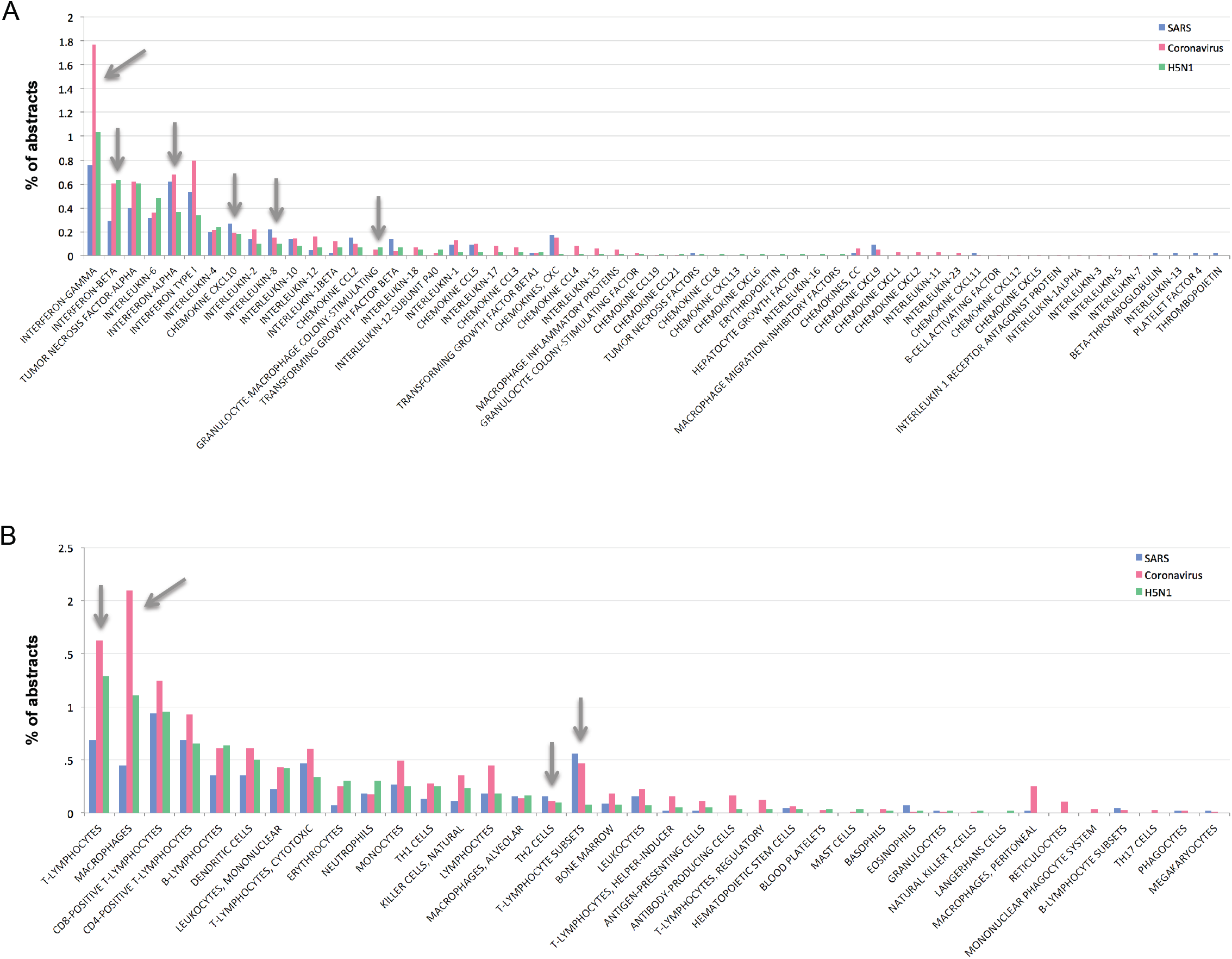
Associations (% of abstracts in respective corpuses) between different entities (MeSh terms) and the three corpuses of PubMed abstracts (those related to SARS, those related to Coronavirus but excluding SARS, and those related to H5N1). A) Different cytokines and chemokines in the three corpuses. B) Different cell types in the three corpuses.

### Immune-related correlations between diseases associated with Coronavirus

Clustering of cytokine-disease co-occurrences in the literature has potentially identified novel relationships between different diseases that are somehow related to Coronavirus. For example, based on these patterns of cytokine associations, the common cold, asthma, and status asthmaticus (a severe form of repetitive asthma attacks) were clustered together (Figure 3). A second cluster included hypertension, nephritis, chorioretinitis and uvetis, while a third interesting cluster included diabetes, hepatitis C and B, panuveitis, pneumonia and pseudorabies. It should be noted however that there was a relatively small number of cytokine-disease co-occurrences in this corpus (when compared for example, to analysis of the entire PubMed database as in [11]).

**Figure 3:**
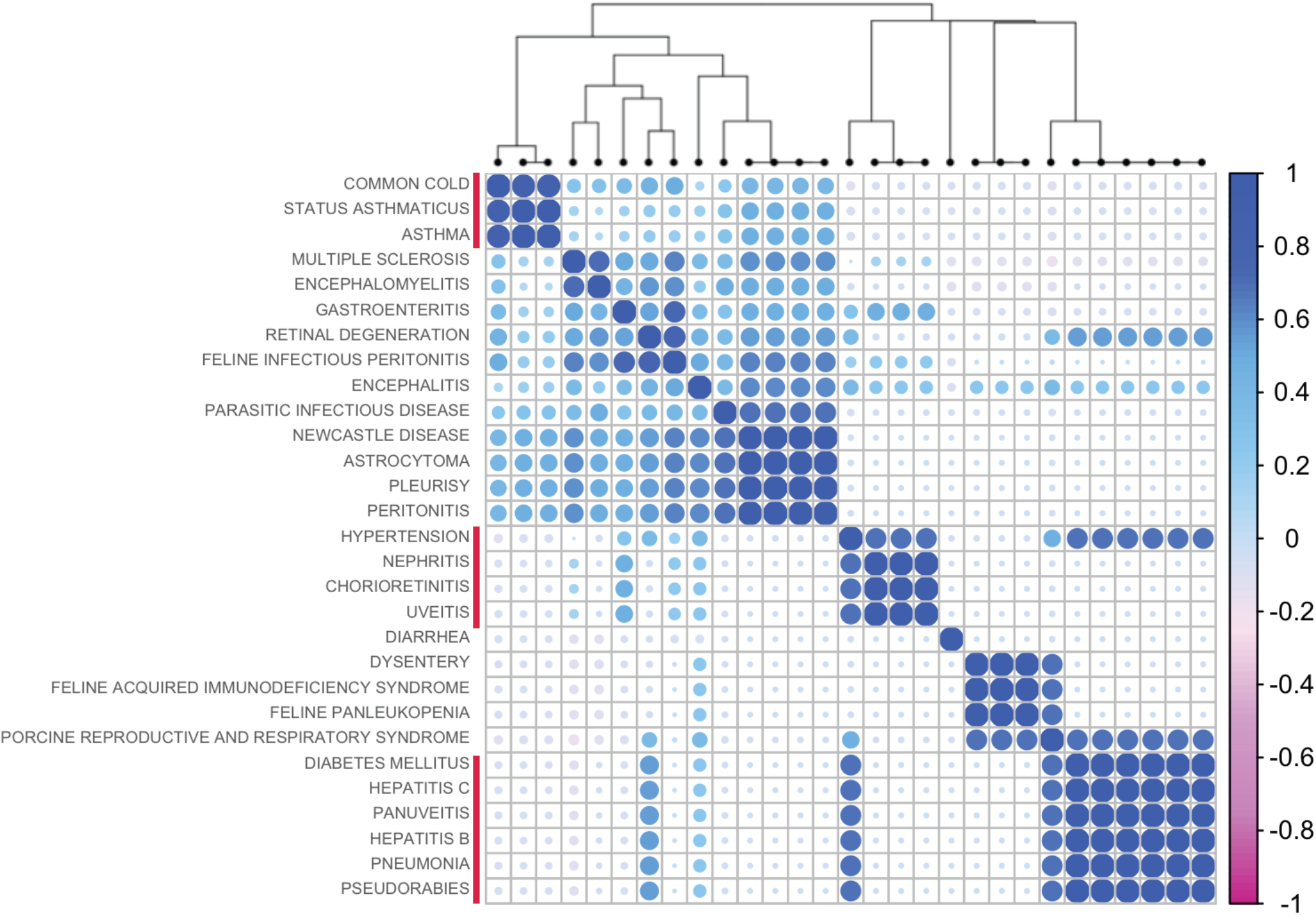
Hierarchical clustering of diseases linked to Coronavirus-related abstracts, based on their patterns of associations with different cytokines.

### Immune interactions in comorbidities of Coronavirus, SARS and H5N1 influenza

Network analysis of the interactions between the different entities captured from PubMed abstracts has identified some interesting and differing sub-networks and connections. For example, in the network generated from the Coronavirus-related associations (Figure 4) different links between cytokines and immune cells and comorbidities or symptoms associated with Coronavirus can be seen. Where asthma and the common cold are directly linked to Interferon-gamma (IFNg), IL-8, IL-2 and IL-5, diarrhea is linked to IL-18.

**Figure 4:**
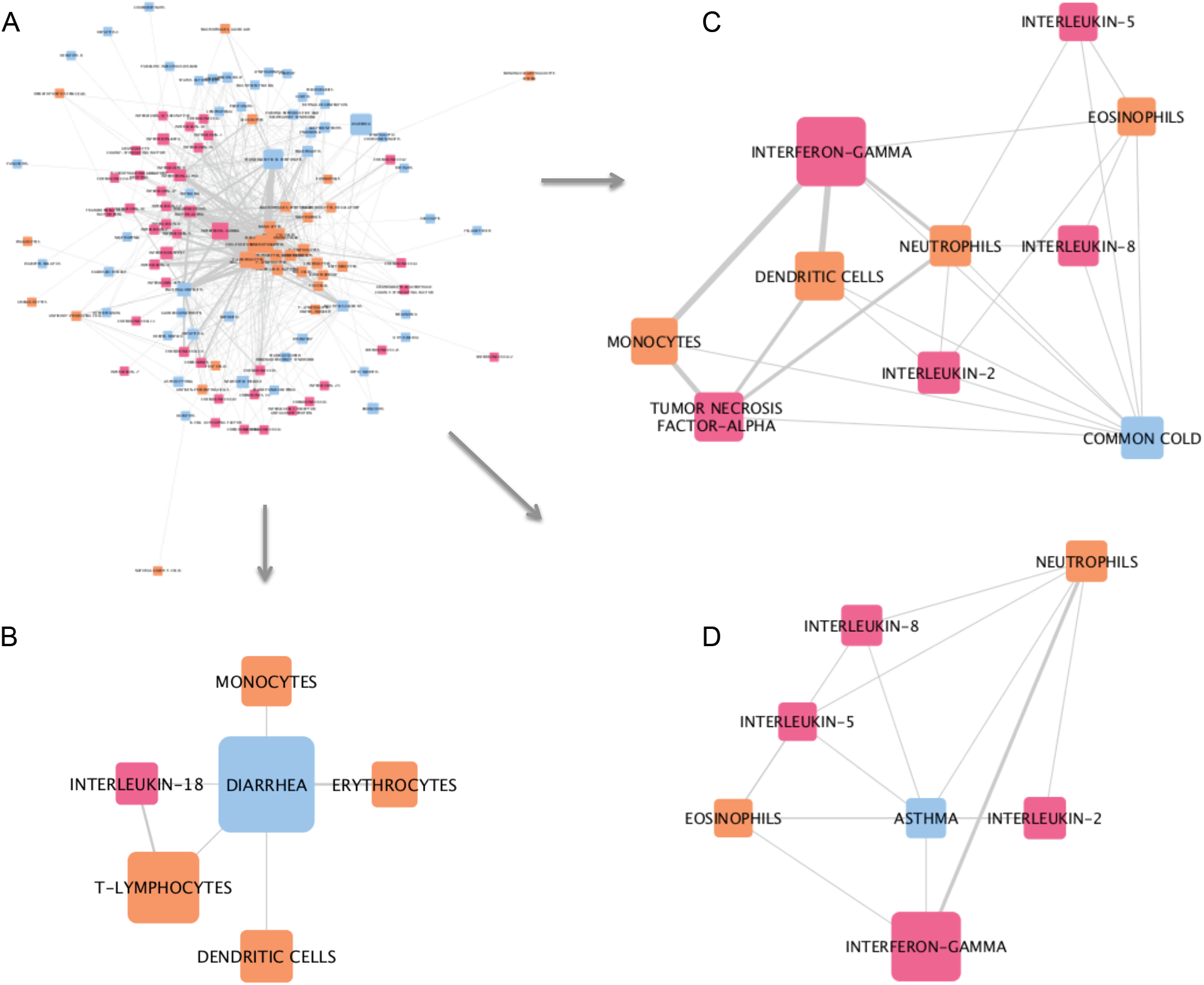
Network analysis of associations captured from MeSH terms in Coronavirus-related PubMed abstracts. A) The whole network. B) Sub-network of first-degree neighbors of the disease node ‘diarrhea’. C) Sub-network of first-degree neighbors of the disease node ‘common cold’. D) Sub-network of first-degree neighbors of the disease node ‘asthma’. Pink nodes represent cytokines, blue nodes represent diseases, and orange nodes represent cell types. The width of edges between nodes corresponds to the number of associations (co-occurrences) between the two entities; the size of the nodes corresponds to the count of that entity within the mined corpus.

Differences in the types of cells included in each sub-network can also be seen. Similarly, in the SARS-related network, we identify differing sub-networks each around a different SARS-related comorbidity (Figure 5); as we did in the H5N1-related network (Figure 6).

**Figure 5:**
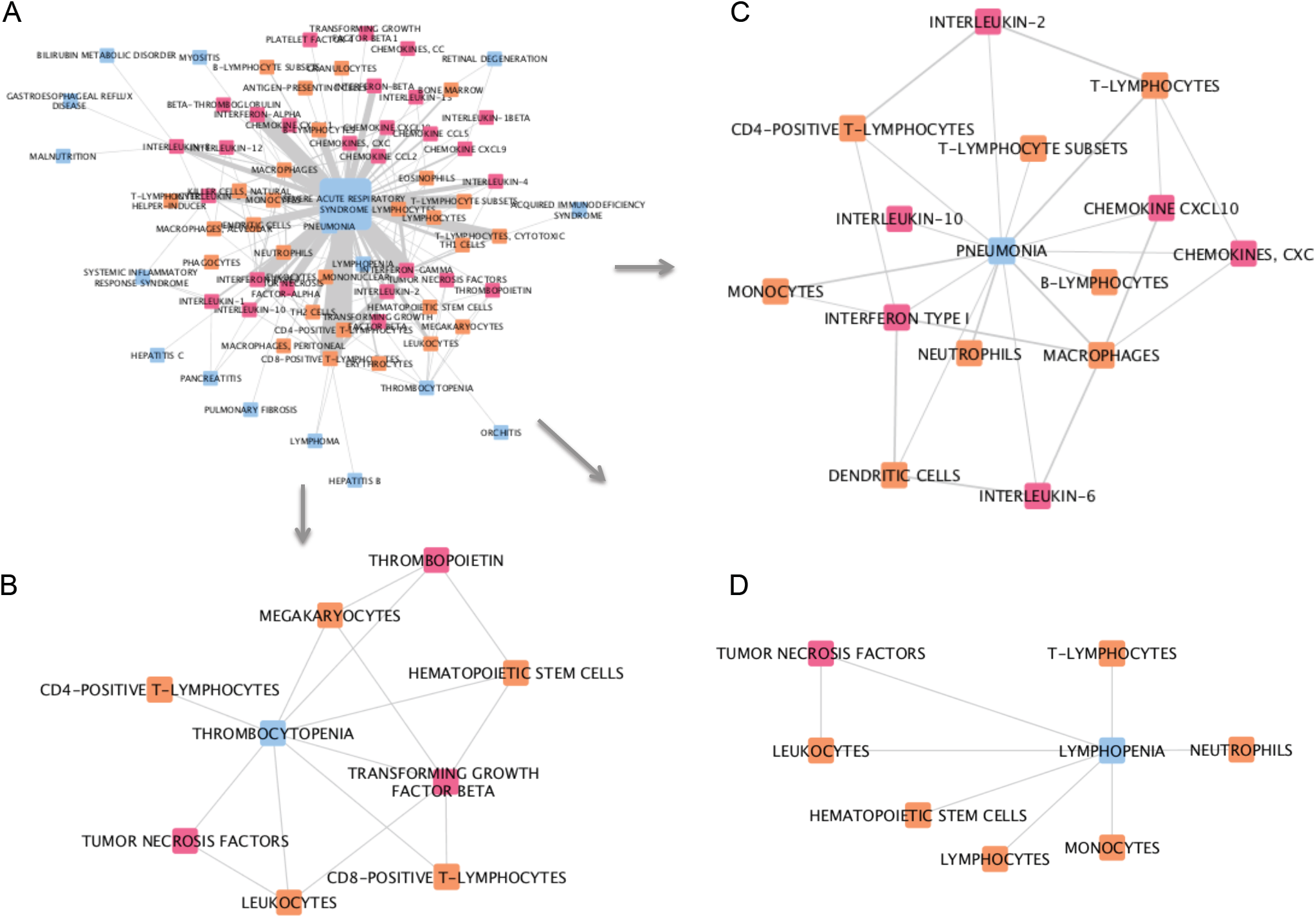
Network analysis of associations captured from MeSH terms in SARS-related PubMed abstracts. A) The whole network. B) Sub-network of first-degree neighbors of the disease node ‘thrombocytopenia’. C) Sub-network of first-degree neighbors of the disease node ‘pneumonia’. D) Sub-network of first-degree neighbors of the disease node ‘lymphopenia’. Pink nodes represent cytokines, blue nodes represent diseases, and orange nodes represent cell types. The width of edges between nodes corresponds to the number of associations (co-occurrences) between the two entities; the size of the nodes corresponds to the count of that entity within the mined corpus.

**Figure 6:**
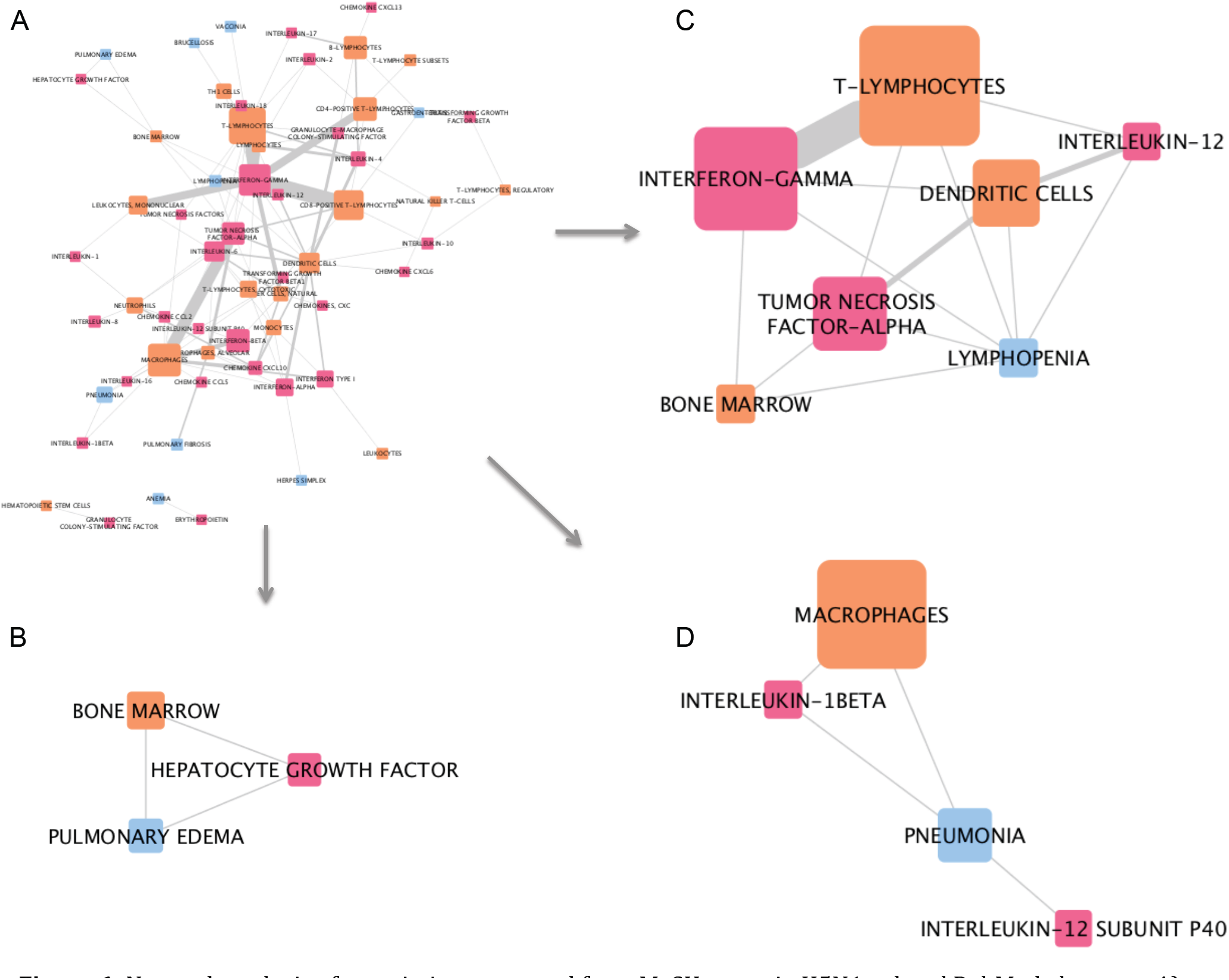
Network analysis of associations captured from MeSH terms in H5N1-related PubMed abstracts. A) The whole network. B) Sub-network of first-degree neighbors of the disease node ‘pulmonary edema’. C) Sub-network of first-degree neighbors of the disease node ‘lymphopenia’. D) Sub-network of first-degree neighbors of the disease node ‘pneumonia’. Pink nodes represent cytokines, blue nodes represent diseases, and orange nodes represent cell types. The width of edges between nodes corresponds to the number of associations (co-occurrences) between the two entities; the size of the nodes corresponds to the count of that entity within the mined corpus.

## DISCUSSION

In this work, we examined possible links of interest between cytokines, immune system cells and diseases in the context of Coronavirus and related infections. By mining information captured within biomedical publications, in the form of MeSH descriptors, and their co-occurrences, we identified associations that may warrant further investigations.

As the literature on SARS-COV-2 develops our approach can be reapplied to identify the strongest, direct links.

In examining the associations between cell types and cytokines, in PubMed abstracts linked to Coronavirus (but not SARS) and those linked to SARS, we found some differences that may point to aetiological differences between SARS-CoV and Coronaviruses more generally. Coronavirus-related abstracts had a significantly higher percentage of associations with Interferon-gamma, Interferon-beta, T-lymphocytes, and macrophages (Figure 2). On the other hand, SARS-related abstracts had higher associations with C-X-C motif chemokine 10 (CXCL10), IL8, and alveolar macrophages; however, these were only marginally higher than the number of associations found in the Coronavirus-related corpus. When comparing these cytokine and cell occurrences to those in H5N1-related abstracts, some patterns are similar to those observed in the Coronavirus-related abstracts; specifically, similar rates of occurrences were observed for IFNb, TNFa, IL-18, and Granulocyte-Macrophage Colony Stimulating Factor (GMCSF), in all cases these were higher than in the SARS-related abstracts. On the other hand H5N1-related abstracts had similar rates of association for monocytes, CD8-positive, and CD4-positive T-lymphocytes to those in SARS, in all cases, lower than those in Coronavirus-related abstracts.

While we cannot rule out that these differences between the three corpuses are partially due to noise, or random variance, it is likely that differences in research focus reflect an underlying cause, and this may reflect biological or clinical differences. The clearest picture of the key features of COVID-19 disease becomes apparent by a consideration of all relevant publications and comparison to related diseases. Here we have shown this is applicable immediately.

Hierarchical clustering of diseases linked to Coronavirus-related abstracts has also identified some interesting groupings of diseases and symptoms associated with Coronavirus infections. For example, the common cold, asthma, and status asthmaticus, are all highly correlated based on their pattern of associations with cytokines within the Coronavirus-related corpus (Figure 3). In a second example, hypertension, nephritis, chorioretinitis and uveitis are all clustered together; and in a third example, diabetes, hepatitis C and B, panuveitis, pseudorabies all cluster with pneumonia. These clusters may suggest similar underlying immunological mechanisms in the context of coronaviruses.

In our third analysis, networks illustrating the connections between cytokines, immune cell types and different diseases, captured from the three abstract corpuses, were generated. Sub-networks, each stemming from a selected disease, illustrate some differences between Coronavirus-, SARS-, and H5N1-related comorbidities and complications. For example, in Coronavirus-related abstracts, diarrhea, one of the reported symptoms of COVID-19, is associated with IL-18, monocytes, T-lymphocytes, dendritic cells and erythrocytes. On the other hand, asthma, a risk factor for adverse outcomes in COVID-19, is linked to IL-8, IL-5, IL-2, IFNg, eosinophils, and neutrophils. This analysis has highlighted some potential targets for therapy. For example, IL-5 is a key growth factor for and activator of eosinophils (both picked up by our research). Eosinophils can be recruited to the lungs, where they have a poorly understood role in health and disease [19]. It has also been reported eosinophil count is a potential marker for COVID-19 patient improvement [20, 21]. The role of eosinophils and IL-5 in this disease certainly requires further investigation. Treatments for asthma that modulate eospinophil action are available; Reslizumab and Mepolizumab are anti-IL-5 antibody therapies that may have potential in treatment of viral respiratory disease, if eosinophils are activated [22, 23]. It is speculative to comment on the role of eosinophils in COVID-19 disease; however in overall consideration of a fast developing literature in a pandemic many possible clinical trial options could be considered. As the literature evolves we suggest the methodologies described here can play a progressive part in rational development of strategies for optimal treatment in an informed environment.

Our approach to capturing immune-related associations from MeSH descriptors is not without limitations. We first assume that co-occurrences of MeSH descriptors within a PubMed record represent a true relationship or dependency; further, some of the associations and co-occurrences were low (i.e. present in only few abstracts). However, our previous investigation into the extent to which MeSH term co-occurrence captures real association has found that at least 70% of co-occurrences of different types of entities (disease, cell type, or cytokine) represent true direct or indirect dependencies, but it is likely to be higher than that [11]. Additionally, patterns of MeSH co-occurrence have shown to capture known medical associations, as well as identify potentially novel ones, thus providing further confidence in the approach. A second limitation is a lack of directionality and type for the associations captured by approach; nevertheless, we show that these mere co-occurrences may still hold valuable information. Finally, since at the time of writing, very few publications directly related to COVID-19 are available, our data mining has had to focus on Coronavirus- and other related lung-damaging diseases as a proxy for COVID-19 Nevertheless, we have created a paradigm for such research which is easy to use and apply, and demonstrated its utility.

## CONCLUSION

Herein, we have identified possible links between immune-related patterns, related co-morbidities, complications and symptoms in the context of Coronavirus and related infections. These associations may have direct implications for COVID-19 and can help focus on potentially useful avenues of future research to understanding of the immune mechanisms underlying COVID-19 and related complications.

## Data Availability

Data on associations mined rom MeSH terms associated with PubMed abstracts are available in Supplementary File 1. All scripts and code used for the mining are available in Geifman et al. JAMIA 2015 (https://doi.org/10.1093/jamia/ocv166).

## ACKNOWLEDGEMENTS

This work was supported by the NIHR Manchester Biomedical research Centre.

NG would like to thank Alice (age 5) for her cooperation, making it possible for this work to be carried out during the COVID-19 pandemic and lockdown.

